# Age-dependent Transcriptional Programs Distinguish Pediatric from Adult Dilated Cardiomyopathy

**DOI:** 10.64898/2026.03.17.26348665

**Authors:** Zoe Leroux, Obed O. Nyarko, Anis Karimpour-Fard, Bonnie Neltner, Matthew L. Stone, Sharon Graw, Luisa Mestroni, Mathew R.G. Taylor, Brian L. Stauffer, Shelley D. Miyamoto, Carmen C. Sucharov

## Abstract

**Background:** Current management of pediatric dilated cardiomyopathy (DCM) in children relies on guideline-directed medical therapy (GDMT) extrapolated from adult heart failure. However, due to small sample size, randomized trials of GDMT agents in children have failed to demonstrate efficacy and mortality benefits seen in adults, suggesting fundamental differences in disease mechanisms. We hypothesized that distinct age-dependent transcriptional programs underlie this therapeutic discordance.

**Methods:** We performed comparative transcriptomic profiling using bulk RNA sequencing on explanted left ventricular tissue from pediatric (n=29) and adult (n=35) DCM patients (adult DCM from previously published data) compared with age-matched non-failing controls (n=22 pediatric, 14 adult). We analyzed differential gene expressions, pathway enrichment across disease etiologies, and the regulation of a conserved 430-gene β1-adrenergic receptor gene signaling network (β1-GSN) known to modulate remodeling in adult heart failure.

**Results:** Transcriptional signatures were profoundly distinct, with only 7.4% of differentially expressed genes shared between adult and pediatric cohorts. Pediatric DCM was characterized by transcriptional reprogramming and the activation of developmental pathways, including WNT/β-catenin and Notch signaling. Conversely, adult DCM hearts were enriched for pathways associated with metabolic dysfunction, mitochondrial deficits, and inflammation. Crucially, while the β1-GSN was desensitized and extensively remodeled in adults, the pathway remained activated in children, with only 4 of 430 network genes showing antithetical regulation.

**Conclusion:** The lack of pathological β-adrenergic remodeling in children could provide a molecular explanation for the lack of clear efficacy of β-blockers in this population. Collectively, these results suggest pediatric DCM represents a biologically distinct disease entity rather than an earlier manifestation of adult heart failure, and future therapeutic strategies must move beyond adult extrapolation to target pediatric-specific pathways.

**Clinical Perspective:** *What Is New?:*

- This comparative transcriptomic analysis demonstrates that pediatric and adult dilated cardiomyopathy possess profoundly distinct biological signatures, with only 7.4% of differentially expressed disease-associated genes shared between the two age groups.
- Unlike adult dilated cardiomyopathy, which is driven by metabolic dysfunction and extensive remodeling of the β1-adrenergic receptor gene signaling network, pediatric disease is uniquely characterized by the activation of developmental pathways such as WNT/β-catenin and Notch signaling without significant pathological β1-adrenergic remodeling.

*What Are the Clinical Implications?:*

- The absence of pathological β-adrenergic remodeling in pediatric hearts provides a molecular explanation for the limited clinical efficacy of β-blocker therapy in this population, underscoring the critical need to develop age-specific precision medicine strategies rather than continuing to extrapolate guideline-directed medical therapies from adult heart failure.

## INTRODUCTION

Functional changes in the β-adrenergic system play an important role in the pathophysiology of heart failure (HF) in adults^1^. Experimental data have extensively defined many of the components involved in the β-adrenergic receptor (β-AR) response in adult HF which resulted in a shift of the clinical treatment paradigm to include β-AR blocker (BB) therapy, which resulted in improvements in morbidity and mortality^2,3^. The treatment of children with HF is largely extrapolated from adult evidence^4^. Despite extrapolation of guideline directed medical therapy (GDMT) from adults to children with DCM, pediatric outcomes have not seen the same dramatic leaps in improvement as has occurred in adults with HF^56^.

GDMT for adults with heart failure is based on the results of large, randomized placebo-controlled clinical trials (RCTs). However, pediatric DCM is a rare disease (0.57–1.13 cases per 100,000 individuals), making it very difficult to perform RCTs to prove efficacy of these adult-based therapies in children. The first RCT performed in children with heart failure studied the non-selective β-blocker, carvedilol. Importantly, although carvedilol has been shown to decrease morbidity and mortality in adults with DCM, the results of the trial were neutral in children^7^. Despite this neutral finding, based on limitations of the trial including a higher-than-expected spontaneous improvement rate in the enrolled participants, treatment with β-blockers remain a Class IIa recommendation for children with HF^8^.

We previously showed, in a small number of hearts, that differences in gene expression in response to DCM are age-specific, which suggests that molecular aspects of the disease are unique in children ^9,10^. Our prior results also highlighted differential adaptation of the β-AR in failing pediatric hearts compared to adult hearts, which could provide a biologic explanation for the limited response to BB in children^9,10^. More recently, a study led by Dr. Michael Bristow identified a beta 1 adrenergic receptor (β_1_-AR) gene signaling network (β_1_-GSN), which constitutes 430 genes dysregulated in mice over-expressing the human β_1_-AR and in adult HF patients^11^. Importantly, these 430 genes exhibited antithetical expression in endomyocardial biopsies from adult HF patients who positively responded to BB treatment when compared to mice over-expressing the human β_1_-AR.

In the current study, we expanded on our prior analysis of the pediatric DCM heart by substantially increasing the number of analyzed DCM and non-failing age-matched healthy control hearts. Through these analyses, we identified age-specific disease pathways altered in the hearts of children that differ from adult DCM hearts. We also evaluated the impact of disease etiology (idiopathic dilated cardiomyopathy (IDC), left ventricular non-compaction (LVNC), or myocarditis) on changes in gene expression. Importantly, we investigated expression of the β_1_-GSN in explanted hearts from pediatric and adult patients with end-stage HF due to non-ischemic DCM. Remarkably, although 47 β_1_-GSN genes were altered in adult DCM hearts, only 4 β_1_-GSN genes were altered in pediatric DCM hearts. Overall, our results suggest that DCM in children is associated with unique age-dependent alterations in molecular pathways, which could be used to identify optimal therapeutic targets in this population.

## METHODS

### Tissue Procurement

Human subjects were males and females younger than 18 years of age, of all races and ethnic background. Patients or their families donated their hearts to the University of Colorado Institutional Review Board (IRB)-approved pediatric transplant tissue bank. Non-failing (NF) control hearts were obtained from donor hearts that could not be used for technical reasons such as blood type mismatch or size. All DCM hearts are from patients without ischemic cardiomyopathy. All patients were evaluated for structural congenital heart disease, primary arrhythmias, neuromuscular disorders or inborn errors of metabolisms, and if present, were excluded from the study. The diagnosis of viral myocarditis is reserved for patients who had evidence of lymphocytic infiltrate associated with myocyte destruction on myocardial biopsy or on histologic evaluation of the explanted heart. The diagnosis of LVNC was made by cardiac imaging echocardiogram and/or cardiac magnetic resonance imaging (MRI) as described in the medical record. At the time of cardiac transplantation or donation, cardiac tissue was rapidly dissected, flash frozen in liquid nitrogen and stored at -80^0^C until further use.

### RNA extraction

RNA was extracted using the Qiagen RNeasy Mini kit with minor modifications. Briefly, 30 mg of tissue from the left ventricle (LV) free wall was homogenized with 600 μl of TRIzol using a Bel-Art homogenizer. After phenol:chloroform extraction, 1 volume of 70% EtOH was added to the clear aqueous phase, mixed and added to the RNeasy column. RNA purification was performed essentially as described by the manufacturer.

### RNA-sequence

RNA-seq of poly-A enriched extracted RNA was performed by the Broad Institute of MIT and sequenced using the Illumina HiSeq 4000 (Illumina, San Diego, CA) platform, for a target depth of ≥40 M 2 × 101 bp paired-end reads. Alignment and quantification of RNA-seq data were performed using the TOPMed RNA-seq Pipeline V2 (CORE yr3) developed by the Broad Institute. The pipeline employs STAR (v2.6.1d) for read alignment to the GRCh38 reference genome, RSEM (v1.3.1) for transcript quantification, and RNA-SeQC (v2.3.3) for quality assessment, using GENCODE v30 gene annotations^12^. Gene expression levels were quantified using transcripts per million (TPM) values generated by RSEM for downstream analysis.

### Analysis and statistics

Comparison between groups were performed using the Wilcoxon rank-sum test. Wilcoxon rank-sum test, *p* and *q*-values were adjusted for multiple testing using the false discovery rate (FDR) method. q-value (adjusted p-values) less than 0.05 were considered statistically significant. Each appropriate statistical test is reported in the figure legends.

### Chi-square analysis

To assess whether the distribution of differentially expressed genes (DEGs) differed between adult and pediatric DCM, chi-square (χ²) analysis was performed on contingency tables made from DEG counts. Genes were grouped by age (adult vs pediatric) and direction of regulation (up- or down-regulated). Statistical significance was determined using Pearson’s chi-square test, with p < 0.05 considered significant.

### Pathway Enrichment Analysis

Gene set enrichment analysis, GO term analysis, Reactome, Wikipathways, KEGG, Biocarta, Hallmark and Ingenuity pathway analysis platforms were performed using DEGs to identify significantly enriched pathways in adult and pediatric DCM compared to non-failing controls. The top significantly enriched pathways for each group were categorized and plotted as figures. Sankey diagrams were generated to visualize the relationship between individual enriched pathways and their corresponding functional categories with link widths proportional to pathway representation within each category.

#### GO Slim Mapping: Source Ontologies and Selection Strategy

Functional categorization was performed using the “goslim_euk_cellular_processes_ribbon” set, a curated subset maintained by the Gene Ontology (GO) Consortium^13^. This set was selected to facilitate a high-level comparative analysis of cellular programs, streamlining granular gene-level data into 61 parent GO terms across three domains: Biological Process (25 terms), Cellular Component (18 terms), and Molecular Function (18 terms). This broad eukaryotic framework is well suited for identifying systemic functional discordance between the pediatric and adult DCM cohorts.

#### Venn Diagram Generation

To compare discordant DEGs between the adult and pediatric DCM groups, we generated area-proportional Venn diagrams using the BioVenn web application ^14^. Adult DEGs were uploaded as ID set X and pediatric DEGs as ID set Y. The “print numbers” option was selected to display absolute counts within each region of the diagram, and the “absolute nrs” setting was used to show the number of overlapping and unique DEGs between groups. Diagram titles and axis labels were entered directly into the interface. For visual clarity, the color for set X (adults) was manually changed to yellow, while the default blue was retained for set Y (pediatrics).

BioVenn allows either absolute numbers or percentages to be displayed, but not at the same time. Because both metrics were required for figure presentation, percentage values were calculated from the BioVenn output and added manually during post-processing in PowerPoint.

To obtain gene lists corresponding to uniquely differentially expressed subsets (FDR < 0.05), we used the Venny 2.1 web application^15^. This tool allowed extraction and download of gene names for each non-overlapping region of the diagram.

#### Canonical Pathway Grouping and Sankey Diagram Construction

To visualize the broader biological impact of DEGs in adult and pediatric DCM, we performed over-representation analysis using the Enrichr web application^16^. Enrichr calculates enrichment using a Fisher’s exact (hypergeometric) test by comparing the number of input genes annotated to each pathway with the total number of genes annotated within each gene-set library, which defines the default background universe. Only DEGs meeting an FDR threshold < 0.05 were included. For co-expression and gene-set expansion features, Enrichr leverages the “All RNA-seq and ChIP-seq Sample and Signature Search” (ARCHS4) resource, which processes human RNA-seq data using the Human Genome Project ^17^, ensuring consistency with current gene annotations.

Enrichment was assessed across the following Enrichr libraries: Reactome Pathways 2024, WikiPathways 2024 Human, KEGG 2021 Human, MSigDB Hallmark 2020, BioCarta 2016, GO Biological Process 2025, GO Cellular Component 2025, and GO Molecular Function 2025.

To create a consistent framework for pathway categorization, enriched terms were manually mapped to higher-order Reactome parent categories using the Reactome Pathway Browser^18^. Pathway names were entered into the search tool, and grouping was based on the closest matching parent term.

Curated pathway groupings were visualized using SankeyMATIC^19^. Diagrams were exported as high-resolution PNG files and manually adjusted for color, spacing, and label clarity.

#### Ingenuity Pathway Analysis (IPA)

Pathway enrichment and network analysis were performed using Ingenuity Pathway Analysis (IPA, Qiagen). Input files contained Ensembl gene identifiers with associated log_2_ fold change, p-value, and false discovery rate (q-value) from bulk RNA-seq differential expression results.

Only genes meeting a significance threshold of q ≤ 0.05 were included in the IPA Expression Core Analysis. This filtering step ensured that pathway enrichment was based exclusively on significantly dysregulated genes. IPA uses the direction of log_2_ fold change to determine pathway activation or inhibition, while the q-value threshold defines the set of genes eligible for enrichment testing.

## RESULTS

### Patient’s characteristics

A summary of subjects’ demographics and clinical characteristics for the pediatric cohort can be found in Table 1. A detailed description of adult patients and NF controls has been published^20^, and a detailed description of pediatric DCM patients and NF controls is depicted in Supplemental Table 1. There were 14 adult NF controls (mean age 56 years, inter-quartile range (IQR) of 14, 78.6% males) of these 21.4% were on BB at the time of collection^20^. There were 35 adult DCM patients (mean age:49 IQR=16, 80% males, average EF 21%, 45.7% were on ventricular assistive device (VAD)) of these 48.6% were on BB at the time of collection^20^. Characteristics of pediatric subjects were as follows: 22 NF controls (mean age:11, IQR=8.6. 63.6% were males) and none were on BB. There were 29 pediatric DCM patients (mean age=5.3, IQR=11.2, 51.7% were males, average EF of 22.7%, 27.6% were on VAD), of these 34.5% on BB. Pediatric DCM patients were further subdivided into IDC (n=23), LVNC (n=3) and myocarditis (n=3)

**Table 1.** A summary of subjects’ demographics and clinical characteristics for the pediatric cohort. **Abbreviations:** DCM = Dilated Cardiomyopathy; NF = Non-failing; IDC = Idiopathic Dilated Cardiomyopathy; LVNC = Left Ventricular Non-Compaction; MC = Myocarditis; FDR = False Discovery Rate; IQR=Interquartile range; EF=Ejection fraction; VAD=Ventricular assistive device.

PEDIATRICS
|  | NF | IDC | LVNC | MC | DCM |
| --- | --- | --- | --- | --- | --- |
| **Number of patients (N)** | 22 | 23 | 3 | 3 | 29 |
| **Age (mean, IQR) (yrs)** | 11, 8.6 | 8.6, 10.3 | 3.97 | 0.67 | 5.3, 11.2 |
| **% population (Male)** | 63.6% | 52.1% | 33% | 66.7% | 51.7% |
| **Average EF %** | 49 | 23 | 31 | 20 | 22.7 |
| **% with VAD** | 0 | 34.8 | 0 | 66.7 | 27.6 |
| **% with Beta-Blockers** | 0 | 43.5 | 0 | 0 | 34.5 |

### Age-Dependent Differences in Gene Expression

To evaluate age-related transcriptional programs in DCM, we compared DEGs in adult and pediatric DCM hearts to their respective age-matched NF controls. As shown in Fig. 1, the majority of genes that change in response to disease were unique to each patient group. We found significant differences in the transcriptional signatures of the adult and pediatric DCM heart (Fig. 1). Only 525 (7.42%) of dysregulated DEGs were commonly shared between the adult and pediatric DCM hearts whereas 3426 (48.43%) DEGs were unique to the pediatric population and 3123 (44.15%) DEGs were unique to the adult population (Fig. 1A)

**Fig. 1.**
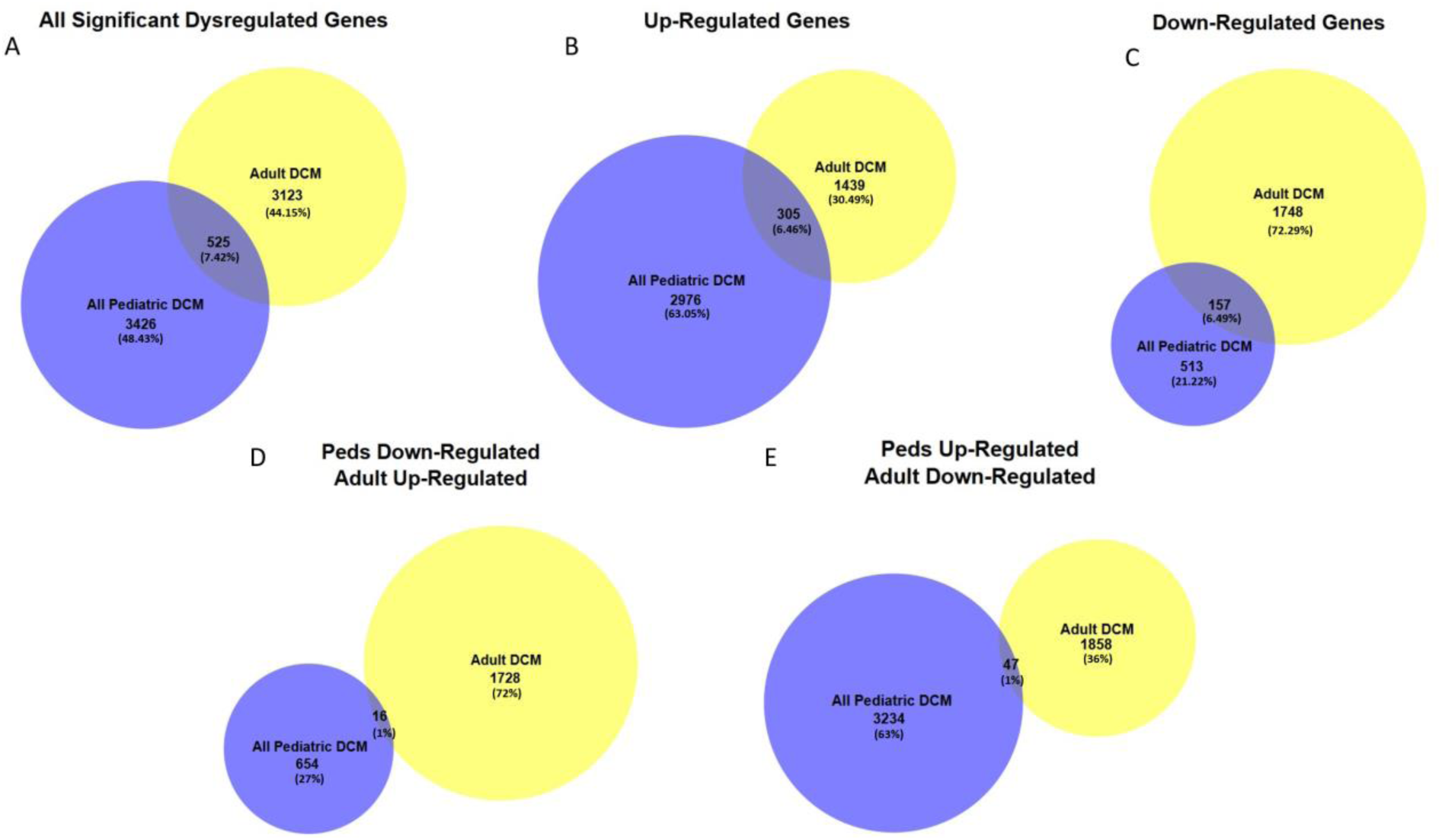
Differential gene expression analysis in adult and pediatric dilated cardiomyopathy (DCM) cohorts compared with age-matched non-failing (NF) controls. Bulk RNA sequencing was performed on left ventricular tissue from adult and pediatric patients with DCM and age-matched NF controls. Pediatric DCM samples included idiopathic DCM (IDC), left ventricular non-compaction (LVNC), and myocarditis (MC). Gene expression values were log₂-transformed, and fold changes were calculated by subtracting the mean NF value from the mean DCM value for each gene. Statistical significance was assessed using Welch’s unequal variance t-test, followed by false discovery rate (FDR) correction to obtain Q-values. Genes with Q ≤ 0.05 were considered significantly dysregulated. (A) Venn diagram representing shared genes between adult and pediatric DCM hearts revealing 525 overlapping dysregulated genes. (B) Venn diagram representing shared upregulated genes between adult and pediatric DCM hearts revealing only 305 overlapping dysregulated genes (C) Venn diagram representing shared downregulated genes between adult and pediatric DCM hearts revealing 157 overlapping dysregulated genes (D) Venn diagram representing shared genes between upregulated adult and downregulated pediatric genes (E) Venn diagram representing shared genes between downregulated adult and upregulated pediatric genes DCM = Dilated Cardiomyopathy; NF = Non-failing; IDC = Idiopathic Dilated Cardiomyopathy; LVNC = Left Ventricular Non-Compaction; MC = Myocarditis; FDR = False Discovery Rate.

Although the proportion of up- (1439 genes) (Fig.1B) and down-regulated genes (1748 genes) (Fig. 1C) was similar in adult DCMs, in children most of the dysregulated genes were up-regulated (2976 upregulated vs 51 downregulated genes). Of the significantly upregulated DEGs, only 305 (6.46%) were common between adult and pediatric DCM hearts, 2976 DEGs (63.05%) were unique to the pediatric DCM hearts and 1439 (30.49%) were unique to adult DCM hearts (Fig. 1B). Similarly, only 157 downregulated DEGs (6.49%) were common between the adult and pediatric DCM hearts with 1748 DEGs (72.29%) unique to the adult DCM and 513 DEGs (21.22%) unique to the pediatric DCM heart (Fig. 1C). Additionally, a small proportion of DEGs were antithetically regulated in pediatric and adult DCM hearts (Fig. 1D and 1E). Our findings highlight distinct transcriptional profiles in pediatric versus adult DCM hearts which suggest age-specific molecular mechanisms of disease.

### Gene Ontology Analyses Highlight Differences in the Biological Response to DCM in Adult and Pediatric Hearts

We next evaluated the biological significance of our findings. GO enrichment analysis of cohort-specific DEGs revealed both shared and divergent functional signatures (Fig. 2). While multiple GO terms in Biological Process (BP), Cellular Component (CC), and Molecular Function (MF) categories were significantly enriched in both cohorts, the magnitude of enrichment and directionality of expression within terms differed substantially between adults and children (Fig. 2A–C). In BP, translation, for example, is significantly altered in adults [-log_10_ (p-value) of 19.70], and marginally significant [-log_10_ (p-value) of 1.556] in children, whereas significance of GO term Signaling is higher [-log_10_ (p-value) of 3.614] in children than adults [-log_10_ (p-value) of 1.641]. Similarly, in CC, Protein-Containing Complex is more significant in adults [-log_10_ (p-value) of 31.590 in adults vs 2.707 in children], and Extracellular Region is more significant in children [-log_10_ (p-value) of 4.967 in children vs 2.236 in adults]. Additionally, the significance of other molecular functions such as transcriptional regulator activity is higher in adults [-log_10_ (p-value) of 32.051 vs 12.969 in children], whereas DNA binding activity is higher in children [-log_10_ (p-value) of 13.305 compared to 9.736 in adults].

**Fig. 2:**
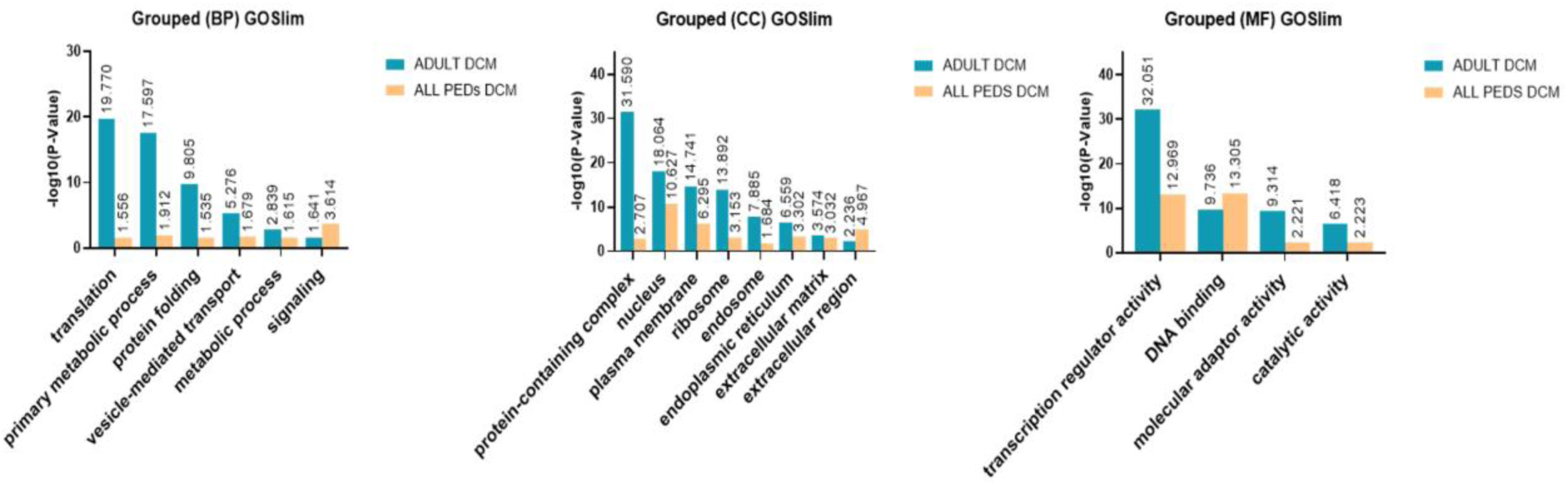
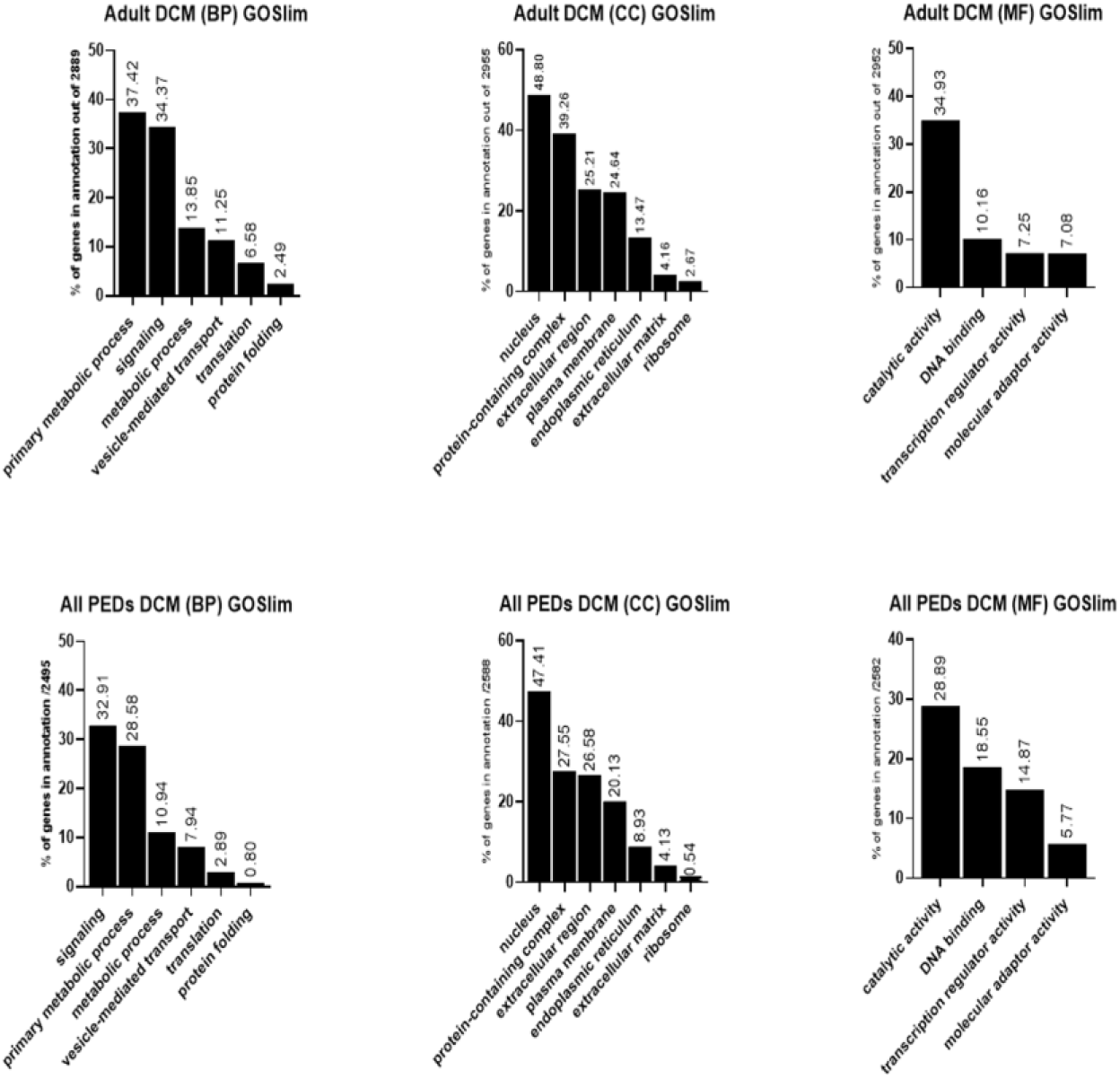
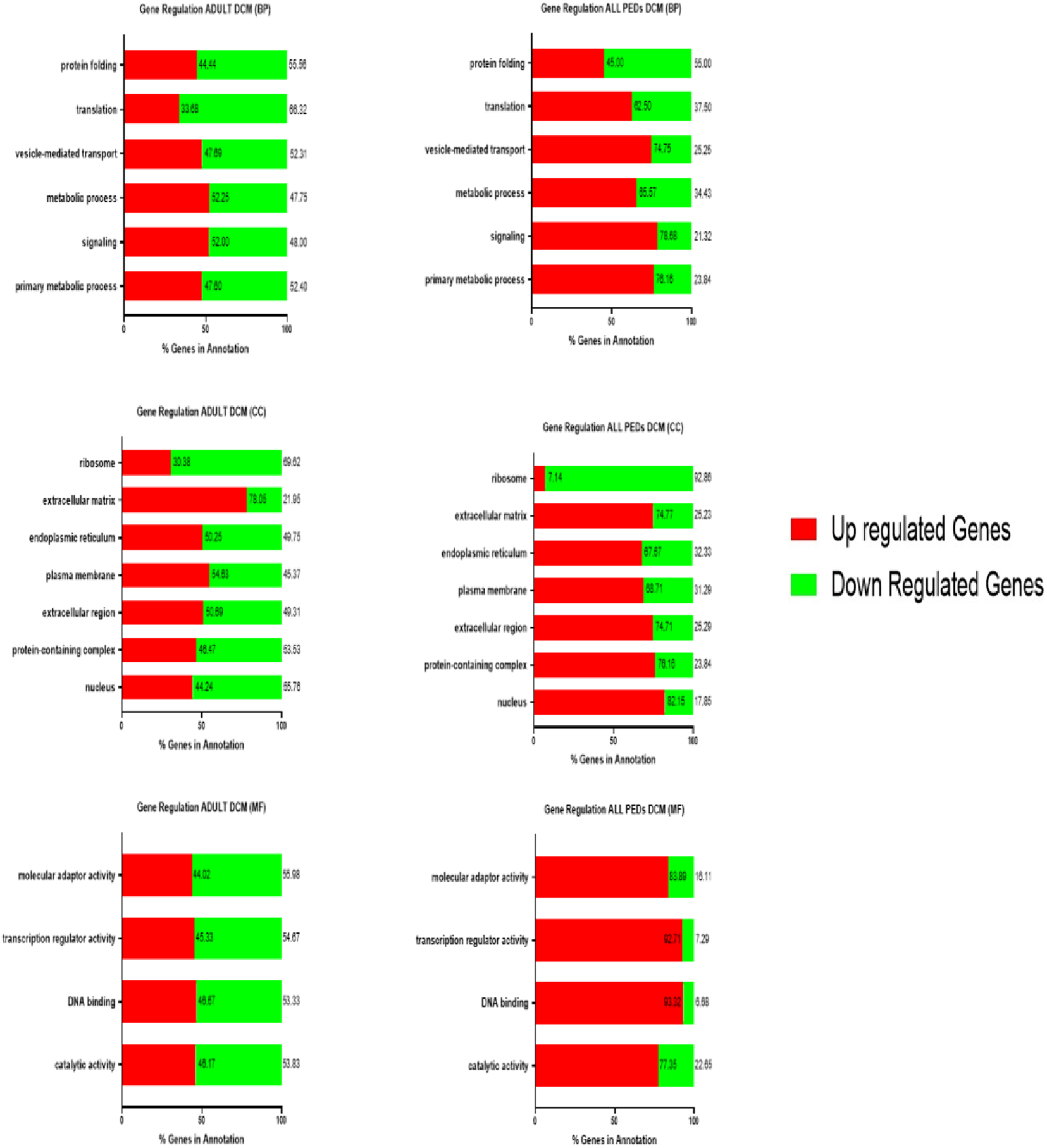
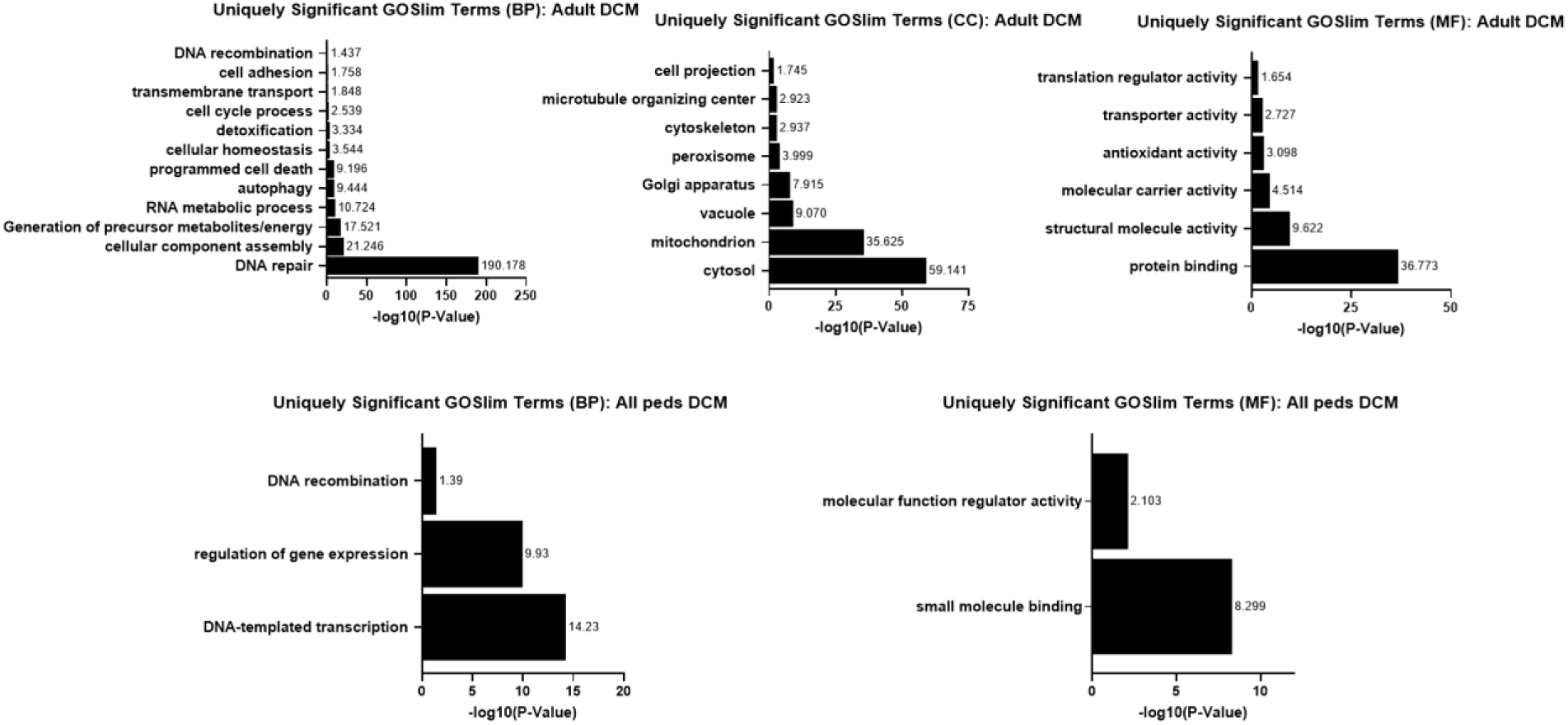
Gene Ontology (GO) Term Enrichment in Adult and Pediatric Dilated Cardiomyopathy (DCM). (A) GO term enrichment analysis was performed on transcriptomic data from adult DCM and pediatric DCM patients using the GOslim pathway analysis tool. The bar graphs display the top enriched GO terms within Biological Processes (BP), Cellular Components (CC), and Molecular Functions (MF). The y-axis represents the negative logarithm of the p-value (-log_10_(p-value)), indicating enrichment significance. (B) For the top enriched GO terms identified in (A), the percentage of genes within the adult DCM and pediatric DCM datasets annotated with these terms is shown. Bar graphs illustrate the proportion of genes associated with specific GO terms for each patient group. (C) For the top enriched GO terms, stacked bar graphs compare the percentage of up-regulated (red) and down-regulated (green) genes between adult DCM and pediatric DCM. (D) Bar graphs display GO slim terms that were significantly enriched in either adult DCM or pediatric DCM, but not in both. The y-axis represents the negative logarithm of the p-value (-log_10_(p-value)), indicating enrichment significance for uniquely significant GO terms.

Fig. 2B depicts the percentage of differentially expressed genes in each of the above GO categories. Although there are similarities in gene distribution, there are dramatic differences in the contribution of up- or down-regulated genes to these processes (Fig. 2C). Analysis of gene expression directionality within GO terms revealed distinct patterns of up- and down-regulation between adult and pediatric DCM. In BP, although the percentage of genes annotated may be similar, such as protein folding (44.44% up-regulated in adults and 45.00% up-regulated in children), in others, there is an inverse distribution of up- and down-regulated genes, such as translation (33.68% up-regulated in adults and 62.50% up-regulated in children). Additionally, differentially expressed genes in vesicle-mediated transport, metabolic process, and signaling in adults are evenly distributed, whereas in children, the majority of altered genes are up-regulated. These differences are also observed for most terms associated with CC and MF. Interestingly, differentially expressed genes associated with ribosome are mostly downregulated (92.86%) in children, whereas they are only 69.92% downregulated in adults. Even more dramatic are the differences observed in MF. In every term, DEGs are balanced in adults, whereas in children, up-regulated genes are the main contributors of predicted alterations. Age-dependent comparisons between GO terms were evaluated by chi-square analysis and are depicted in Supplemental Table 2.

Fig. 2D depicts significant GO Slim term analyses. These highlight distinct biological processes enriched in adult versus pediatric DCM. BP in adults are highly associated with cellular processes, whereas in children, they are transcriptional/nuclear processes. There are no significant changes in GO Slim CC in children. MF in pediatric DCM likely relates to binding of molecules to proteins. In adults, protein binding seems to relate to protein-protein interactions that modulate protein function.

The BP, CC and MFs of IDC, LVNC and myocarditis further emphasize the uniqueness of these components even with respect to the different etiologies of DCM (Supp. Fig. 1). Interestingly, myocarditis is highly enriched in processes associated with cell death, DNA repair and replication, whereas IDC is associated with cellular homeostasis and metabolic processes. Very few processes are unique to LVNC, which may indicate that alterations in response to LVNC are more diffuse.

Interestingly in Fig. 3, Gene Ontology terms suggest DEGs in adults (Fig. 3a) are highly associated with alterations in proteins, including translation and ubiquitination, respiration, including several mitochondria-associated terms, extracellular matrix remodeling, and inflammation. In children (Fig. 3b), although some metabolic and inflammatory pathways are predicted to be altered, differentiation, development, and structural organization processes are highly significant pathways. Additionally, the only CC predicted to be altered in children are associated with components of extracellular matrix remodeling, whereas mitochondrial alterations are over-represented in adults. Interestingly, molecular functions in children are highly associated with epigenetic mechanisms, whereas in adults, protein translation/regulation is prevalent. (Fig. 3).

**Fig. 3:**
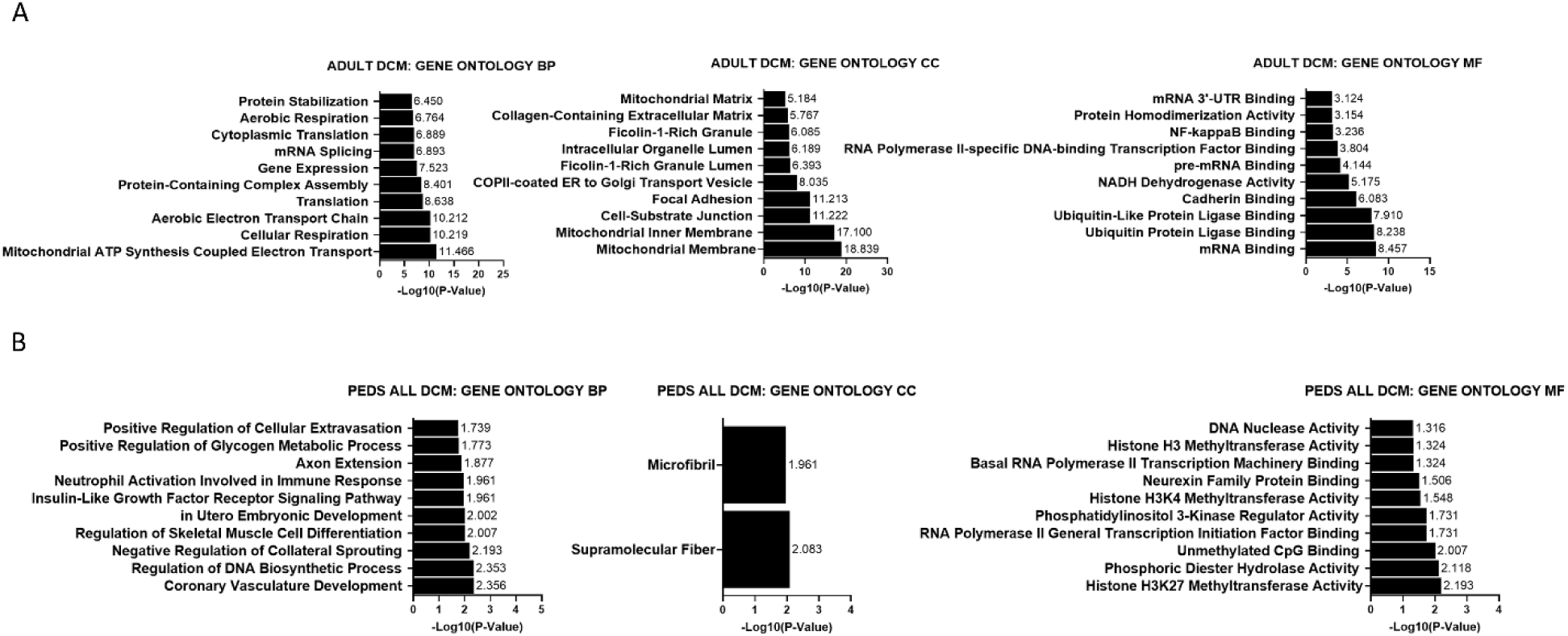
Top 10 Gene Ontology (GO) terms significantly enriched in cardiac RNA sequencing data from adult and pediatric dilated cardiomyopathy (DCM) samples. (A) Gene enrichment analysis was performed using Enrichr, and the top 10 most significant GO terms (p-value ≤ 0.05) are displayed for Biological Process (BP) (left), Cellular Component (CC)(middle), and Molecular Function (MF) (right) ontologies for adult DCM. The x-axis represents the -log_10_(P-value), indicating the significance of the enrichment. Higher values correspond to more significant enrichment of the GO term within the differentially expressed gene lists. (B) Gene enrichment analysis was performed using Enrichr, and the top 10 most significant GO terms (p-value ≤ 0.05) are displayed for Biological Process (BP) (left), Cellular Component (CC)(middle), and Molecular Function (MF) (right) ontologies for pediatric DCM. The x-axis represents the -log_10_(P-value), indicating the significance of the enrichment. Higher values correspond to more significant enrichment of the GO term within the differentially expressed gene lists.

### Gene set enrichment analysis and GO terms also highlight distinct molecular signatures and biological pathways in adult and pediatric DCM hearts

Using curated genes from the six major pathway databases (Reactome, WikiPathways, BioCarta, Hallmark, KEGG, and Ingenuity Pathway Analysis), we visualized the top pathways using Sankey diagrams and categorized them into broader functional groups. In adult DCM (Fig. 4a), metabolism-related categories were prevalent. In contrast, pediatric DCM (Fig. 4b) cellular processes, such as signal transduction, vesicle-mediated transport and cell cycle, were enriched. (Fig. 4).

**Fig. 4:**
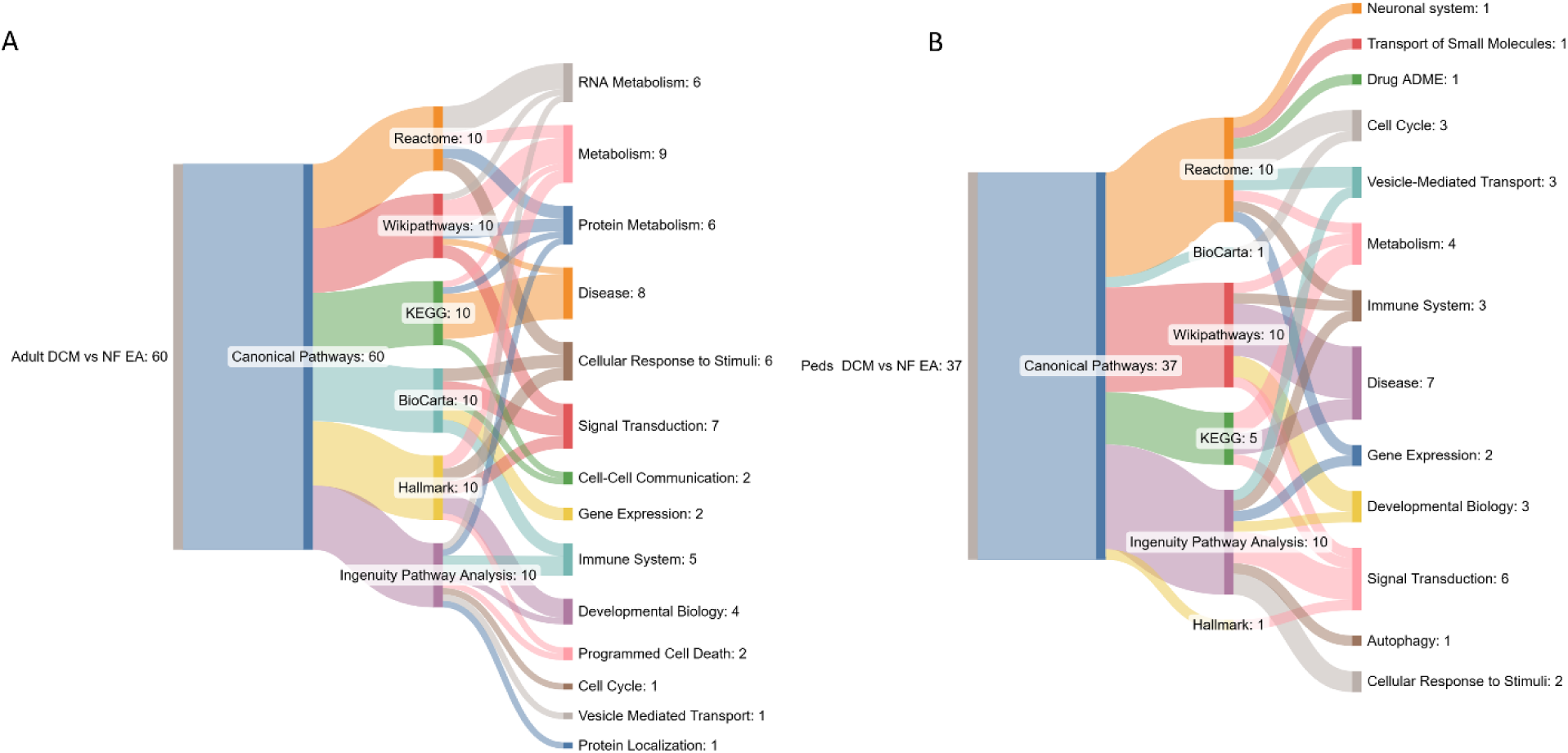
Gene set enrichment analysis (GSEA) of canonical pathways in adult and pediatric idiopathic dilated cardiomyopathy (DCM) compared with non-failing (NF) controls. (A) The Sankey diagrams illustrate the top 10 most statistically enriched canonical pathways identified by GSEA in adult DCM versus NF hearts, derived from six major pathway databases: Reactome, WikiPathways, BioCarta, Hallmark, KEGG, and Ingenuity Pathway Analysis. Each pathway is classified into broader functional categories. In adult DCM, the most enriched categories were metabolism (35%), disease-related pathways (13%), and signal transduction (12%). Functional categories uniquely enriched in adults included cell–cell communication, programmed cell death, and protein localization. (B) The Sankey diagrams illustrate the top 10 most statistically enriched canonical pathways identified by GSEA in pediatric DCM versus NF hearts, derived from six major pathway databases: Reactome, WikiPathways, BioCarta, Hallmark, KEGG, and Ingenuity Pathway Analysis. Each pathway is classified into broader functional categories. In pediatric DCM, the predominant categories were disease (19%), signal transduction (16%), and metabolism (11%). Functional categories uniquely enriched in pediatric-specific pathways included autophagy, neuronal system, transport of small molecules, and drug absorption, distribution, metabolism, and excretion (ADME).

GSEA pathway analysis also identified unique patterns of pathway activation in adult vs pediatric DCM. We observed a predicted upregulation of Epithelial to Mesenchymal Transition (EMT), TGF-β signaling, and TNF-α signaling in adult DCM. On the other hand, we found activation of development-associated pathways, such as hedgehog signaling, WNT/β-catenin and Notch signaling, in pediatric DCM. There was also a unique downregulation of immune pathways such as inflammatory signals, TNF-α, and IL6-JAK-STAT3 signaling in pediatric DCM (Fig.5A, B). We also used GSEA to better understand differences in disease etiology. Interestingly, WNT/β-catenin signaling is upregulated in IDC and myocarditis but downregulated in LVNC. Inflammatory response is upregulated in LVNC but downregulated in IDC (Fig.5C-E). Myocarditis is enriched in cell cycle-associated processes, transcriptional regulation, and response to interferon. EMT is uniquely up-regulated in pediatric IDC. These data underscore both age-dependent and etiology-dependent heterogeneity in DCM molecular pathways

**Fig. 5.**
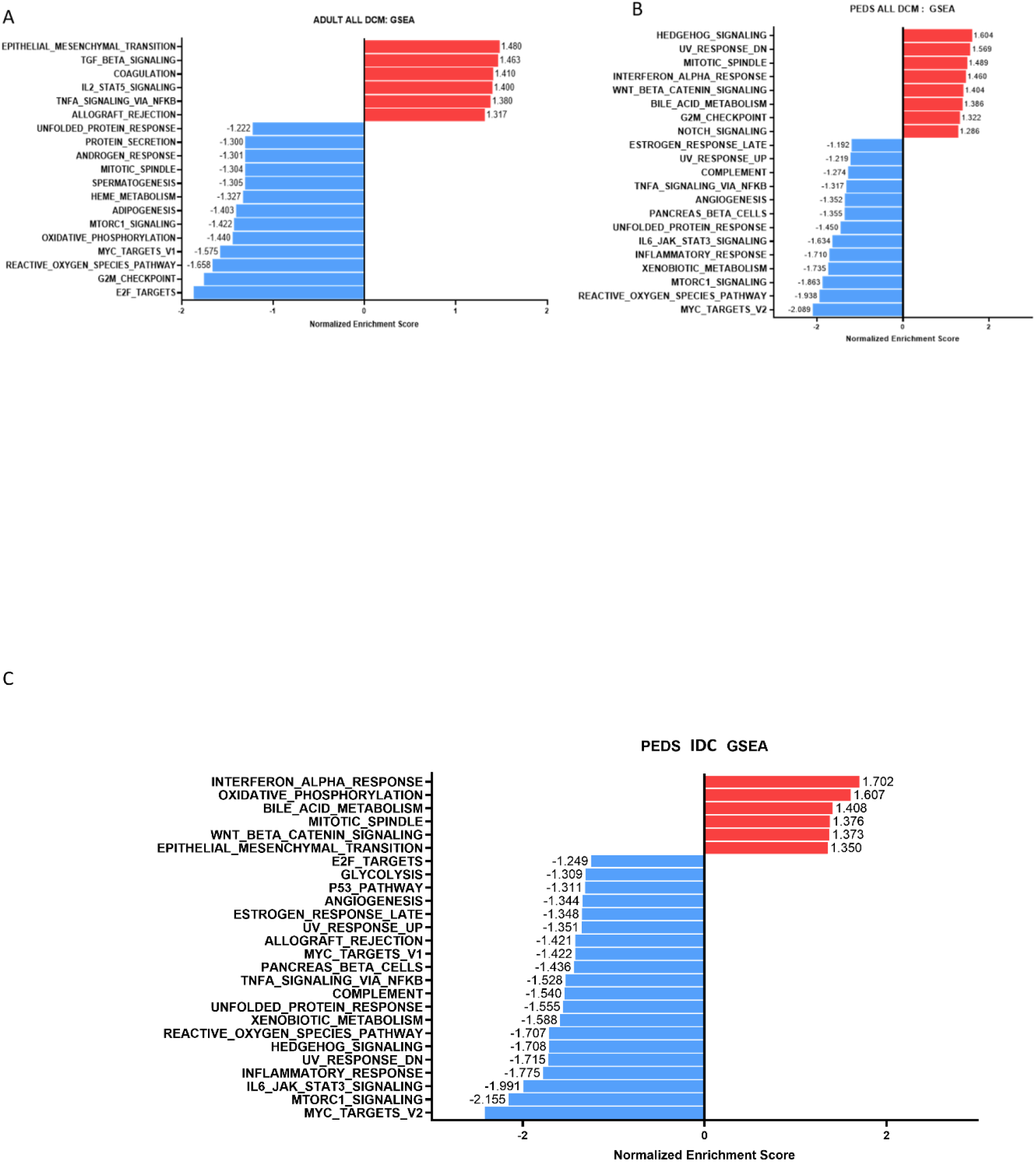
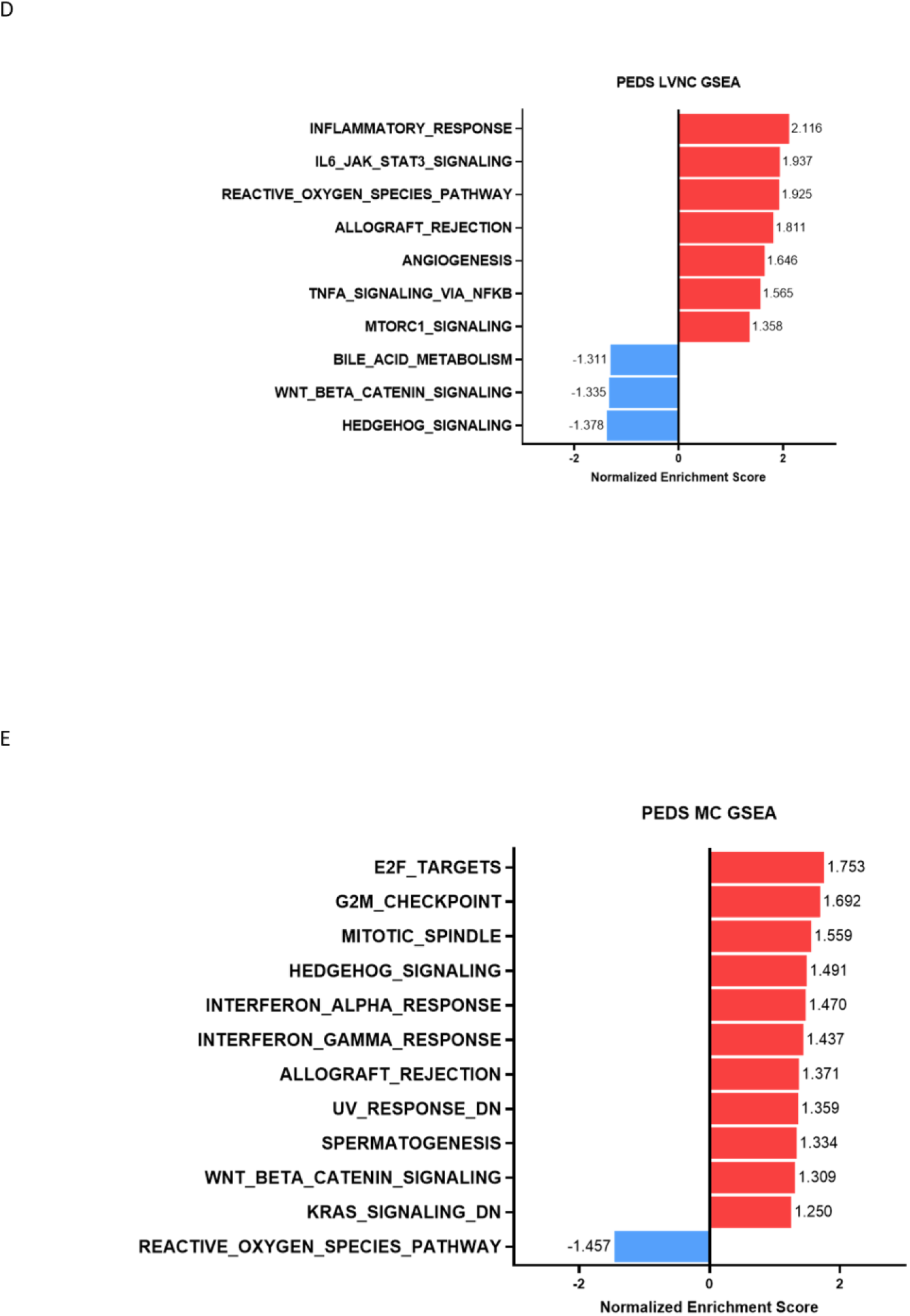
Gene set enrichment analysis highlights distinct biological pathways in adult and pediatric DCM hearts. (A) Gene set enrichment analysis using DEGs in adult DCM hearts compared to non-failing controls showing upregulated (red) and downregulated pathways(blue) (B) Gene set enrichment analysis using DEGs in pediatric DCM hearts compared to non-failing controls showing upregulated (red) and downregulated pathways(blue) (C) Gene set enrichment analysis using DEGs in pediatric idiopathic DCM(IDC) hearts compared to non-failing controls showing upregulated (red) and downregulated pathways(blue) (D) Gene set enrichment analysis using DEGs in pediatric left ventricular non-compaction (LVNC) hearts compared to non-failing controls showing upregulated (red) and downregulated pathways(blue) (E) Gene set enrichment analysis using DEGs in pediatric myocarditis hearts compared to non-failing controls showing upregulated (red) and downregulated pathways(blue)

### Unique transcriptional signatures in the β_1_-adrenergic receptor gene signaling network in adult vs pediatric DCM

Given the observed differences in gene expression profiles, enrichment in biological processes and resultant predictive pathways, we next investigated whether the distinct transcriptional signatures extend to phenotypic alterations and responses to key cardiac signaling systems and therapies. We focused on the β_1_-AR system, a key and highly effective therapeutic target in adult heart failure. We have previously shown that there exists a highly conserved β_1_-AR responsive gene signaling network (β_1_-GSN) in adults with DCM and highlighted its role in modulating pathologic ventricular remodeling^11,21^.

Of the 430 β_1_-GSN, we identified 47 genes antithetically regulated in end stage adult DCM hearts and in the hearts of patients who responded to BB. In contrast, in children with DCM, only four of the 430 genes in the β_1_-GSN were antithetically regulated (Fig. 6). These genes include ENTPD6, MYH6, TMED3 and TIMP1, which encode diverse proteins related to cardiac function and cellular metabolism. Interestingly, and in support of these analyses, IPA analysis of the adult and pediatric DCM heart shows a profound difference in predicted activation of the β-AR signaling pathway in children and adults, with a predicted pathway inhibition in adults (z-score=-1.941) and a predicted pathway activation in children (z-score=3.162) (Table 2). These findings are in agreement with pathway desensitization observed in adults, and suggest this pathway is still activated in children with end-stage HF.

**Fig. 6:**
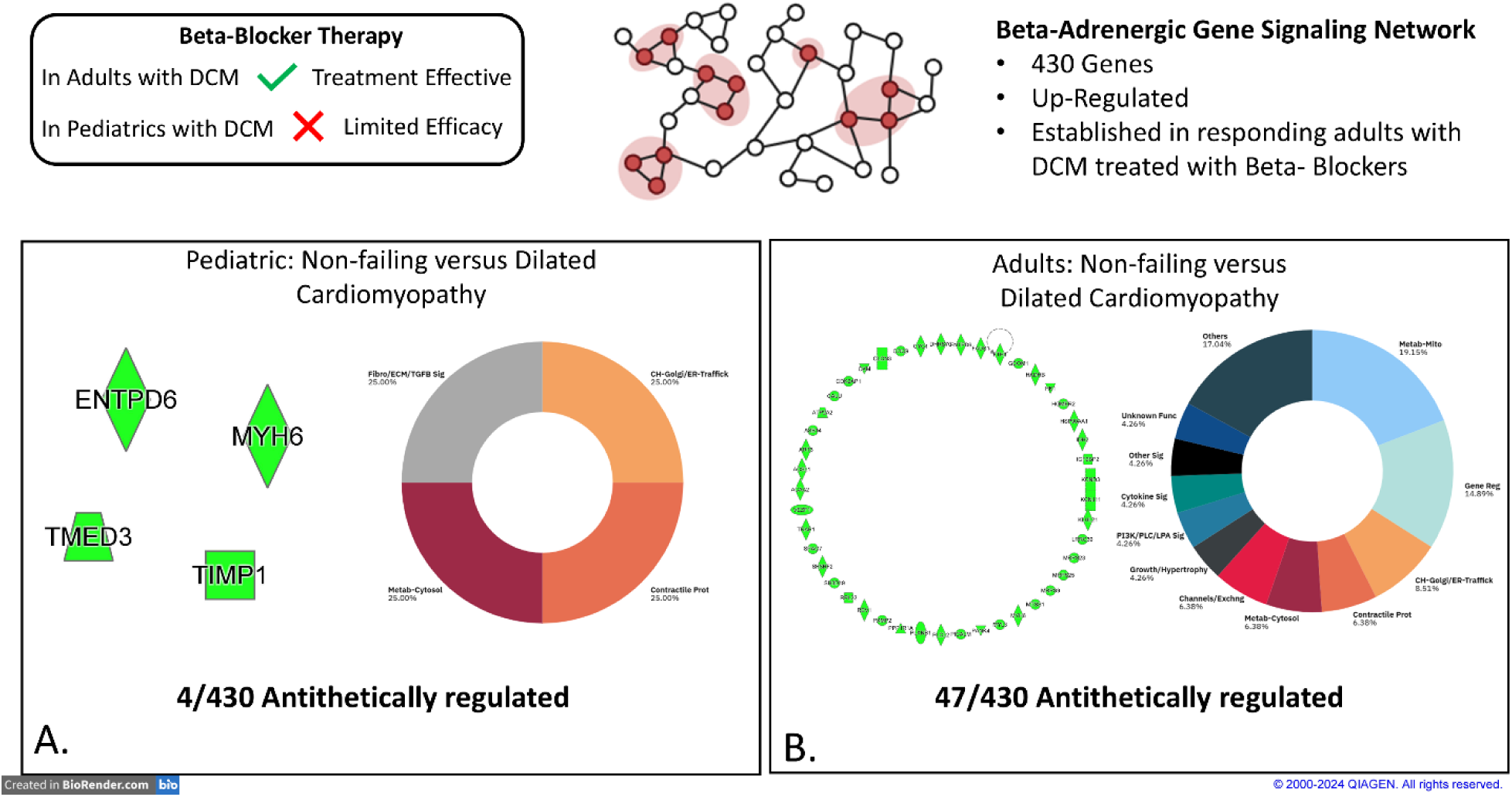
Transcriptional response of the beta-adrenergic signaling pathway in pediatric dilated cardiomyopathy (DCM) versus adult dilated cardiomyopathy (DCM). (A) This Illustrates the expression profile in pediatric DCM, where only 4 genes (1%) showed antithetical regulation. The four antithetically regulated genes in pediatric DCM fell into four categories: fibroblast/ECM/TGF-β signaling, cellular homeostasis/Golgi/ER trafficking, cytosolic metabolism, and contractile proteins (B) This shows 47 genes (11%) were oppositely regulated in adult DCM, suggesting a more robust transcriptional response. Functional annotation of these 47 genes in adults revealed involvement in diverse biological processes including: metabolism/mitochondrial, cytosolic, and sarcoplasmic reticulum; gene regulation; contractile proteins; signaling pathways such as PI3K/PLC/LPA, adrenergic/β-adrenergic/PKA, non-β-adrenergic, calcium handling, cytokines, and other signaling categories; cellular homeostasis; Golgi/ER trafficking, mitochondrial integrity, and peroxisomal integrity; apoptosis; extracellular matrix and fibroblast/TGF-β signaling; growth and hypertrophy regulation; ion channels and solute exchangers; and unknown functions. DCM = Dilated Cardiomyopathy; IDC = Idiopathic Dilated Cardiomyopathy; Metab-Mito = Mitochondrial Metabolism; Metab-Cytosol = Cytosolic Metabolism; Metab-SR = Sarcoplasmic Reticulum Metabolism; Gene Reg = Gene Regulation; Contractile Prot = Contractile Proteins; PI3K/PLC/LPA Sig = Phosphoinositide 3-Kinase/Phospholipase C/Lysophosphatidic Acid Signaling; Adr/BAR/PKA Sig = Adrenergic/β-Adrenergic/Protein Kinase A Signaling; Ca²⁺ Handling = Calcium Handling; Cytokine Sig = Cytokine Signaling; Other Sig = Other Signaling Pathways; CH-Golgi/ER-Traffick = Cellular Homeostasis–Golgi/Endoplasmic Reticulum Trafficking; CH-Mito Integrity = Cellular Homeostasis–Mitochondrial Integrity; CH-Peroxi Integrity = Cellular Homeostasis–Peroxisomal Integrity; Fibro/ECM/TGFB Sig = Fibroblast/Extracellular Matrix/Transforming Growth Factor-β Signaling; Growth/Hypertrophy = Growth and Hypertrophy Signaling; Channels/Exchng = Channels and Solute Exchangers; Unknown Func = Unknown Function.

**Table 2.** IPA analysis of the adult and pediatric DCM heart with predicted pathway activation and inhibitions for adult and pediatric hearts. **Abbreviations:** DCM = Dilated Cardiomyopathy; NF = Non-failing; IDC = Idiopathic Dilated Cardiomyopathy; LVNC = Left Ventricular Non-Compaction; MC = Myocarditis.

**Ingenuity Canonical Pathway: Cardiac β-adrenergic Signaling**
| Pathology Group | $-\log(p\text{-value})$ | Ratio | z-score |
| --- | --- | --- | --- |
| B-Blocker Responders | 4.14 | 0.0476 | 2.449 |
| Adult DCM | 0.49 | 0.155 | -1.941 |
| PEDs ALL DCM | 0.00 | 0.133 | 3.162 |
| PEDs IDC | 0.00 | 0.0181 | NA |
| PEDs LVNC | 0.00 | 0.0061 | NA |
| PEDs MC | 0.623 | 0.0976 | 1.890 |

## DISCUSSION

In this study, we provide a comprehensive comparative transcriptomic analysis of adult and pediatric DCM, revealing profound age and etiology-dependent differences in gene expression, biological pathway enrichment, and signaling responses. Despite sharing a common clinical phenotype of DCM, our data demonstrate that the biological, cellular and molecular processes underlying adult and pediatric DCM are distinct, with limited overlap in DEGs. These findings suggest that DCM in children and adults may represent mechanistically unique disease entities. The unique transcriptional landscape of pediatric compared to adult DCM further expands to key therapeutic cardiac signaling systems such as the β-adrenergic system.

### Distinct Age-Specific Transcriptional Profiles

Only about 7% of DEGs were shared between adult and pediatric DCM, emphasizing the limited commonality in the transcriptional response to disease across age groups. This difference may reflect heightened developmental plasticity in pediatric DCM, in which stress responses preferentially activate transcriptional programs that support growth, differentiation, and remodeling^22^. Cellular stress activates transcriptional reprogramming, a process essential for sustaining homeostasis in pathological states^23^. In pediatric DCM, transcriptional regulation predominated, with enrichment of pathways associated with cardiac development. These transcriptional pathways have important roles in cardiomyocyte development, maturation, differentiation and specification^24,25^, and may reflect an activated, highly plastic transcriptional state. Such activation could represent the myocardium’s attempt to adapt to injury through reactivation of fetal-like developmental processes or could be an intrinsic cause of disease. Exploring the role of these pathways in pediatric DCM and targeting them may uncover an age-specific therapeutic avenue.

By contrast, adult DCM was characterized by enrichment of pathways associated with translation and protein regulation, including ribosomal biogenesis, ubiquitination, mitochondrial oxidative phosphorylation, and proteostasis networks. These findings are consistent with impaired protein homeostasis, metabolic dysfunction, and cellular senescence, which are hallmarks of chronic and multi-factorial disease progression in adult heart failure^26^. For example, ribosomal and mitochondrial pathways were among the most significantly activated pathways in adults, consistent with cardiac bioenergetics energy deficits in adult heart failure^27,28^. In children, mitochondrial or metabolic dysfunction may occur without strong transcriptional reprogramming because the disease progresses relatively quickly.

These differences suggest that pediatric DCM represents a pathology dominated by transcriptional reprogramming and activation of developmental pathways, whereas adult DCM reflects chronic maladaptive changes in protein synthesis and turnover, metabolism, and cellular senescence. This dichotomy may explain the faster clinical disease progression observed in children in contrast to the chronic and cumulative disease course typical of adult DCM.

### Distinct Biological Processes and Pathways in adult vs pediatric DCM

Gene ontology and GSEA further emphasize the distinct biological mechanisms underlying DCM in adults and children. Importantly, the activation/repression of these signaling pathways differed substantially by age. Pediatric DCM was enriched for transcriptional, developmental, and structural organization processes, including pathways such as Hedgehog, WNT/β-catenin and Notch signaling. In the heart, prior studies have implicated hedgehog signaling in the regulation of cardiogenesis including cardiomyocyte formation and specification^29^, cardiomyocyte proliferation and maturation ^30^ and coronary vascular development^31^. Similarly, WNT/β-catenin and Notch signaling also play critical roles in the development and homeostasis of the heart and in maintaining the cardiac gene network^25,32^. We have shown that WNT/β-catenin and Notch signaling are activated in pediatric DCM hearts and in the hearts of a novel model of pediatric DCM that recapitulates important aspects of pediatric DCM^11,33,34^. We and others have suggested targeting WNT/β-catenin and Notch signaling as a novel therapeutic target in pediatric DCM^25,33^.

Conversely, adult DCM hearts demonstrated enrichment in metabolic pathways, mitochondrial processes, and protein turnover, aligning with known impairments in energy metabolism and proteostasis in failing adult hearts^35^. Pathways such as TNF-α signaling, TGF-β signaling, and extracellular matrix remodeling were also highly upregulated, in keeping with the chronic inflammatory and fibrotic responses that characterize the adult disease course, which differs from pediatric DCM^36,37^.

This difference further emphasizes the unique characteristics of pediatric DCM when compared to the well-known phenotypic changes observed in the adult heart^38^, including minimal fibrosis^39^, lack of cardiomyocyte hypertrophy^21^, unique β−AR remodeling^40^, and a diminished immune cell response^41^. This divergence in molecular mechanisms suggests that age-specific therapeutic strategies focusing on transcriptional and developmental pathways in children, and metabolic function and fibrotic pathways in adults may be critical to improving outcomes in both populations.

### β1-Adrenergic Signaling Networks are distinct in adult vs pediatric DCM

Given the central role of β-adrenergic signaling in heart failure pathophysiology and therapy, we investigated the β_1_-GSN in the pediatric DCM transcriptome. Our key analysis highlights a markedly attenuated remodeling of this signaling axis in children. β_1_-AR pathway desensitization is a hallmark of adult HF. While this predicted desensitization was observed in adult hearts, in children, this pathway is predicted to be activated, suggesting that β_1_-AR desensitization in pediatric hearts does not impact downstream signaling pathways affected in adults. Although previous studies have demonstrated downregulation of β_1_-adrenergic receptor density and classical desensitization in pediatric DCM^10^, our transcriptomic pathway analysis predicted activation of β_1_-AR related signaling in children. These findings are not contradictory when considering that receptor abundance and downstream transcriptional responses are regulated at distinct cellular levels^42^. Pediatric hearts may engage compensatory signaling via β_2_-AR^43,44^, or crosstalk with developmental transcriptional networks^45^, resulting in sustained or heightened downstream activity of genes associated with β-adrenergic signaling despite the reduction of β_1_-AR density. Additionally, it is possible that the β_1_-GSN in children has not yet been activated due to the faster disease progression in children. Together, these data highlight age-specific adrenergic adaptations that may influence both disease progression and therapeutic response.

In adults, we previously defined a conserved β_1_-GSN composed of 430 gene members that are dynamically regulated in response to β-blocker therapy or β_1_-AR stimulation, with 47 network genes significantly altered in adult DCM. This network encompasses genes involved in contractility, metabolism, and extracellular matrix regulation, highlighting its broad role in driving adverse remodeling. We have previously shown that both β_1_- and β_2_-AR were downregulated in pediatric DCM, while in adults only β_1_-AR was reduced with preserved β_2_-AR expression ^9,10^. Collectively, these results suggest a differential adaptation of the β-AR signaling system in pediatric compared to adult DCM and that the β_1_-AR pathway may not be a dominant driver of pediatric disease, which may explain, at the molecular level, why β-blocker therapy, a cornerstone of adult heart failure management, has not definitively improved outcomes in children^46^.

The identification of antithetically regulated genes in the pediatric β1-GSN further highlights the divergence in age-specific biology. For example, MYH6, encoding α-myosin heavy chain, is developmentally regulated and typically downregulated in HF regardless of etiology, so likely not a β-AR specific change^47^. While expressed in many tissues, Ectonucleoside triphosphate diphosphohydrolase 6 (ENTPD6) is highly expressed in the heart^48^. ENTPD6 is a soluble member of the ectonucleoside triphosphate diphosphohydrolase family released from injured tissues that can influence extracellular signaling and cardiac remodeling ^49^. ENTPD6 has not been extensively studied, and its role in β-AR signaling is largely unknown. Similarly, transmembrane p24 protein transport domain containing 3 (TMED3), a member of the transmembrane p24 domain family, regulates vesicular transport intracellularly and has been well studied for its role in various cancers via WNT signaling but has not yet been studied in the context of HF^50^. On the other hand, Tissue Inhibitor of Metalloproteinases 1 (TIMP1) has been well studied in the context of HF. TIMP1 is a member of a family of inhibitors of the matrix metalloproteinases that play a role in the degradation of the extracellular matrix (ECM) ^51^. TIMP1 promotes myocardial fibrosis and is associated with myocardial remodeling^52^. TIMP1 expression is activated by β-AR stimulation ^53^ and can also promote β-AR-mediated apoptosis in the heart^54^. These findings are consistent with our prior work and work by others^10,11,55^, which demonstrated developmental stage-dependent changes in transcriptional responses to β-adrenergic signaling ^53^. In summary, the lack of β-adrenergic remodeling in pediatric DCM provides a potential molecular explanation for the limited efficacy of β-blocker therapy in children.

### Clinical and Therapeutic Implications

Our findings provide an important perspective when considering the clinical management of pediatric DCM. First, we suggest that pediatric DCM is not simply a “smaller scale” version of the adult disease but instead reflects distinct transcriptional and signaling mechanisms. Second, age-specific differences in β1-AR signaling raise the possibility that some HF therapies optimized for adults may be suboptimal in children. Indeed, the modest clinical benefit of β-blockers in pediatric trials may reflect the limited involvement of β1-GSN pathways in pediatric disease biology. Additionally, pathways uniquely enriched in pediatric DCM including WNT/β-catenin and Notch signaling pathways represent potential novel therapeutic targets specific to pediatric DCM patients.

### Limitations and Future Directions

The current study focuses on gene expression changes in the heart. Pathway analyses are predictive of the consequences of these changes, but no functional measurements have been evaluated. These analyses were performed in end-stage explanted hearts, making it difficult to determine if the observed changes represent cause or consequence of the disease. Future *in vivo* studies are necessary to investigate the functional significance of these changes and their impact over time.

## CONCLUSION

In summary, we demonstrated that adult and pediatric DCM exhibit profoundly distinct transcriptional and biological signatures, including differences in cardiac pathways such as β1-adrenergic signaling, metabolism, development, Hedgehog, Notch and WNT signaling. These findings underscore the need to consider biological age as a modifier of the drivers of DCM pathophysiology and responsiveness to treatment strategies. Our results provide a premise for the development of not only age- but etiology-specific therapeutic approaches that more effectively target the unique molecular mechanisms driving disease in pediatric and adults with DCM.

## Acknowledgments

Molecular data for the Trans-Omics in Precision Medicine (TOPMed) program was supported by the National Heart, Lung and Blood Institute (NHLBI). RNASeq for “NHLBI TOPMed: Trans-Omic in Precision Congestive Heart Failure (TOPCHeF) (phs002038; NHLBI 1X01HL139403-01)” was performed at the Broad Institute (HHSN268201600034I). Core support including centralized genomic read mapping and genotype calling, along with variant quality metrics and filtering were provided by the TOPMed Informatics Research Center (3R01HL-117626-02S1; contract HHSN268201800002I). Core support including phenotype harmonization, data management, sample-identity QC, and general program coordination were provided by the TOPMed Data Coordinating Center (R01HL-120393; U01HL-120393; contract HHSN268201800001I). We gratefully acknowledge the studies and participants who provided biological samples and data for TOPMed. We would also like to acknowledge the Heart Transplant Team at Children’s Hospital Colorado.

## Data Availability

The datasets supporting the current study including raw, processed and metadata of RNA sequencing will be made publicly available in NCBI GEOdatabase upon acceptance of the manuscript for publication.

## Author contributions

ZL and OON contributed equally to this work and were responsible for data analysis, data interpretation, figure preparation, and drafting of the manuscript. AKF contributed to bioinformatic analyses, data analysis, and interpretation of transcriptomic datasets. BN processed tissue samples, extracted RNA and performed quality control. MLS contributed to acquisition of pediatric clinical samples and interpretation of pediatric clinical data. SG, LM and MRGT are PIs of the TOPmed grant which supported RNA-sequencing of all samples. BLS, SDM and CCS conceived and supervised the study, were responsible for study design, and were involved in data interpretation, and manuscript preparation.

All authors reviewed the manuscript and approved the final version for publication.

## REFERENCES

1. Bernstein D, Fajardo G, Zhao M. THE ROLE OF β-ADRENERGIC RECEPTORS IN HEART FAILURE: DIFFERENTIAL REGULATION OF CARDIOTOXICITY AND CARDIOPROTECTION. Prog. Pediatr. Cardiol. [Internet]. 2011 [cited 2025 Dec 8];31:35. Available from: https://pmc.ncbi.nlm.nih.gov/articles/PMC3135901/

2. Lymperopoulos A, Rengo G, Koch WJ. The Adrenergic Nervous System in Heart Failure: Pathophysiology and Therapy. Circ. Res. [Internet]. 2013 [cited 2025 Dec 8];113:10.1161/CIRCRESAHA.113.300308. Available from: https://pmc.ncbi.nlm.nih.gov/articles/PMC3843360/

3. Parichatikanond W, Duangrat R, Kurose H, Mangmool S, Parichatikanond W, Duangrat R, Kurose H, Mangmool S. Regulation of β-Adrenergic Receptors in the Heart: A Review on Emerging Therapeutic Strategies for Heart Failure. Cells 2024, Vol. 13, [Internet]. 2024 [cited 2025 Dec 8];13. Available from: https://www.mdpi.com/2073-4409/13/20/1674

4. Irving C, Azeka E, Adorisio R, Blume ED, Bogle C, Chubb H, Conway J, Cousino MK, Edelson J, Ford K, et al. The International Society for Heart and Lung Transplantation Guidelines for the Management of Pediatric Heart Failure (Update From 2014). Journal of Heart and Lung Transplantation [Internet]. 2025 [cited 2025 Dec 7];1–51. Available from: 10.1016/j.healun.2025.06.003

5. Loss KL, Shaddy RE, Kantor PF. Recent and Upcoming Drug Therapies for Pediatric Heart Failure. Front. Pediatr. [Internet]. 2021 [cited 2022 Sep 25];9:11. Available from: /pmc/articles/PMC8632454/

6. Merlo M, Cannatà A, Pio Loco C, Stolfo D, Barbati G, Artico J, Gentile P, De Paris V, Ramani F, Zecchin M, et al. Contemporary survival trends and aetiological characterization in non-ischaemic dilated cardiomyopathy. Eur. J. Heart Fail. [Internet]. 2020 [cited 2025 Dec 7];22:1111–1121. Available from: https://pubmed.ncbi.nlm.nih.gov/32452075/

7. Shaddy RE, Curtin EL, Sower B, Tani LY, Burr J, LaSalle B, Boucek MM, Mahony L, Hsu DT, Pahl E, et al. The pediatric randomized carvedilol trial in children with chronic heart failure: Rationale and design. Am. Heart J. [Internet]. 2002 [cited 2025 Dec 7];144:383–389. Available from: https://pubmed.ncbi.nlm.nih.gov/12228773/

8. Irving C, Azeka E, Adorisio R, Blume ED, Bogle C, Chubb H, Conway J, Cousino MK, Edelson J, Ford K, et al. The International Society for Heart and Lung Transplantation Guidelines for the Management of Pediatric Heart Failure (Update From 2014). The Journal of Heart and Lung Transplantation [Internet]. 2025 [cited 2026 Mar 1];44:e21–e71. Available from: https://www.jhltonline.org/action/showFullText?pii=S1053249825020303

9. Miyamoto SD, Sucharov CC, Woulfe KC. Differential Response to Heart Failure Medications in Children. Prog. Pediatr. Cardiol. [Internet]. 2018 [cited 2025 Dec 7];49:27. Available from: https://pmc.ncbi.nlm.nih.gov/articles/PMC6023416/

10. Miyamoto SD, Stauffer BL, Nakano S, Sobus R, Nunley K, Nelson P, Sucharov CC. Beta-adrenergic adaptation in paediatric idiopathic dilated cardiomyopathy. Eur. Heart J. [Internet]. 2014 [cited 2025 Sep 28];35:33–41. Available from: https://pubmed.ncbi.nlm.nih.gov/22843448/

11. Tatman PD, Kao DP, Chatfield KC, Carroll IA, Wagner JA, Jonas ER, Sucharov CC, David Port J, Lowes BD, Minobe WA, et al. An extensive β1-adrenergic receptor gene signaling network regulates molecular remodeling in dilated cardiomyopathies. JCI Insight [Internet]. 2023 [cited 2025 Dec 7];8:e169720. Available from: https://pmc.ncbi.nlm.nih.gov/articles/PMC10543724/

12. TOPMed_RNAseq_pipeline_flowchart_COREyr3.

13. Gene Ontology Resource [Internet]. [cited 2026 Jan 6];Available from: https://geneontology.org/

14. Hulsen T, de Vlieg J, Alkema W. BioVenn - A web application for the comparison and visualization of biological lists using area-proportional Venn diagrams. BMC Genomics. 2008;9.

15. Venny 2.1.0 [Internet]. [cited 2026 Jan 6];Available from: https://bioinfogp.cnb.csic.es/tools/venny/

16. Kuleshov M V., Jones MR, Rouillard AD, Fernandez NF, Duan Q, Wang Z, Koplev S, Jenkins SL, Jagodnik KM, Lachmann A, et al. Enrichr: a comprehensive gene set enrichment analysis web server 2016 update. Nucleic Acids Res. 2016;44:W90–W97.

17. Homo sapiens genome assembly GRCh38 - NCBI - NLM [Internet]. [cited 2026 Jan 6];Available from: https://www.ncbi.nlm.nih.gov/datasets/genome/GCF_000001405.26/

18. Home - Reactome Pathway Database [Internet]. [cited 2026 Jan 6];Available from: https://reactome.org/

19. SankeyMATIC: Make Beautiful Flow Diagrams [Internet]. [cited 2026 Jan 6];Available from: https://sankeymatic.com/

20. Sweet ME, Cocciolo A, Slavov D, Jones KL, Sweet JR, Graw SL, Reece TB, Ambardekar A V., Bristow MR, Mestroni L, et al. Transcriptome analysis of human heart failure reveals dysregulated cell adhesion in dilated cardiomyopathy and activated immune pathways in ischemic heart failure. BMC Genomics [Internet]. 2018 [cited 2025 Dec 16];19. Available from: https://pubmed.ncbi.nlm.nih.gov/30419824/

21. Tatman PD, Woulfe KC, Karimpour-Fard A, Jeffrey DA, Jaggers J, Cleveland JC, Nunley K, Taylor MRG, Miyamoto SD, Stauffer BL, et al. Pediatric dilated cardiomyopathy hearts display a unique gene expression profile. JCI Insight [Internet]. 2017 [cited 2023 Jul 28];2. Available from: /pmc/articles/PMC5518568/

22. de Magalhães-Barbosa MC, Prata-Barbosa A, da Cunha AJLA. Toxic stress, epigenetics and child development. J. Pediatr. (Rio. J). [Internet]. 2021 [cited 2025 Sep 28];98:S13. Available from: https://pmc.ncbi.nlm.nih.gov/articles/PMC9510910/

23. Himanen S V., Sistonen L. New insights into transcriptional reprogramming during cellular stress. J. Cell Sci. [Internet]. 2019 [cited 2025 Sep 28];132. Available from: https://pubmed.ncbi.nlm.nih.gov/31676663/

24. Arnhold V, Boos J, Lanvers-Kaminsky C. Targeting hedgehog signaling pathway in pediatric tumors: in vitro evaluation of SMO and GLI inhibitors. Cancer Chemother. Pharmacol. [Internet]. 2016 [cited 2025 Sep 28];77:495–505. Available from: https://pubmed.ncbi.nlm.nih.gov/26781311/

25. Azhdari M, zur Hausen A. Wnt/β-catenin and notch signaling pathways in cardiovascular disease: Mechanisms and therapeutics approaches. Pharmacol. Res. [Internet]. 2025 [cited 2025 Sep 28];211. Available from: https://pubmed.ncbi.nlm.nih.gov/39725339/

26. Li Z, Zhao H, Wang J. Metabolism and Chronic Inflammation: The Links Between Chronic Heart Failure and Comorbidities. Front. Cardiovasc. Med. [Internet]. 2021 [cited 2025 Sep 28];8:650278. Available from: https://pmc.ncbi.nlm.nih.gov/articles/PMC8131678/

27. Lopaschuk GD, Karwi QG, Tian R, Wende AR, Abel ED. Cardiac Energy Metabolism in Heart Failure. Circ. Res. [Internet]. 2021 [cited 2025 Sep 28];128:1487–1513. Available from: https://www.ahajournals.org/doi/10.1161/CIRCRESAHA.121.318241

28. Jana S, Zhang H, Lopaschuk GD, Freed DH, Sergi C, Kantor PF, Oudit GY, Kassiri Z. Disparate Remodeling of the Extracellular Matrix and Proteoglycans in Failing Pediatric Versus Adult Hearts. J. Am. Heart Assoc. [Internet]. 2018 [cited 2025 Apr 24];7. Available from: https://pubmed.ncbi.nlm.nih.gov/30371322/

29. Gibb N, Lavery DL, Hoppler S. Sfrp1 promotes cardiomyocyte differentiation in Xenopus via negative-feedback regulation of Wnt signalling. Development (Cambridge) [Internet]. 2013 [cited 2023 Jul 29];140:1537–1549. Available from: https://pubmed.ncbi.nlm.nih.gov/23482489/

30. Waldron CJ, Kelly LA, Stan N, Kawakami Y, Abrahante JE, Magli A, Ogle BM, Singh BN. The HH-GLI2-CKS1B network regulates the proliferation-to-maturation transition of cardiomyocytes. Stem Cells Transl. Med. [Internet]. 2024 [cited 2025 Sep 28];13:678–692. Available from: https://pubmed.ncbi.nlm.nih.gov/38761090/

31. Lavine KJ, Kovacs A, Ornitz DM. Hedgehog signaling is critical for maintenance of the adult coronary vasculature in mice. J. Clin. Invest. [Internet]. 2008 [cited 2025 Sep 28];118:2404. Available from: https://pmc.ncbi.nlm.nih.gov/articles/PMC2430494/

32. Wang Y, Lu P, Wu B, Morrow BE, Zhou B. NOTCH maintains developmental cardiac gene network through WNT5A. J. Mol. Cell. Cardiol. 2018;125:98–105.

33. Nyarko OO, Rausch E, Goff JRH, Karimpour-Fard A, Conard CS, Hernandez-Lagunas L, Burns MPA, Peña B, Miyamoto SD, Stauffer BL, et al. A novel Notch and WNT signaling mechanism contribute to pediatric DCM: a pathway to new therapeutics. bioRxiv [Internet]. 2025 [cited 2025 Sep 28];Available from: https://pubmed.ncbi.nlm.nih.gov/40501561/

34. Moran HR, Nyarko OO, Garfield AL, O’Rourke R, Ching RCK, Gray R, Cobb TM, Riemslagh FW, Lovely C Ben, Peña B, et al. The pericardium forms as a distinct structure during heart formation. Nat. Commun. [Internet]. 2025 [cited 2025 Oct 30];16:8566. Available from: https://pmc.ncbi.nlm.nih.gov/articles/PMC12480573/

35. Morciano G, Vitto VAM, Bouhamida E, Giorgi C, Pinton P. Mitochondrial Bioenergetics and Dynamism in the Failing Heart. [Internet]. 2021 [cited 2025 Sep 28];11:436. Available from: https://pmc.ncbi.nlm.nih.gov/articles/PMC8151847/

36. Rao W, Li D, Zhang Q, Liu T, Gu Z, Huang L, Dai J, Wang J, Hou X. Complex regulation of cardiac fibrosis: insights from immune cells and signaling pathways. J. Transl. Med. [Internet]. 2025 [cited 2025 Sep 28];23:242. Available from: https://pmc.ncbi.nlm.nih.gov/articles/PMC11869728/

37. Biernacka A, Dobaczewski M, Frangogiannis NG. TGF-β signaling in fibrosis. Growth Factors [Internet]. 2011 [cited 2025 Sep 28];29:196. Available from: https://pmc.ncbi.nlm.nih.gov/articles/PMC4408550/

38. Lee TM, Hsu DT, Kantor P, Towbin JA, Ware SM, Colan SD, Chung WK, Jefferies JL, Rossano JW, Castleberry CD, et al. Pediatric Cardiomyopathies. Circ. Res. [Internet]. 2017 [cited 2023 Jul 28];121:855–873. Available from: https://www.ahajournals.org/doi/abs/10.1161/CIRCRESAHA.116.309386

39. Woulfe KC, Siomos AK, Nguyen H, SooHoo M, Galambos C, Stauffer BL, Sucharov C, Miyamoto S. Fibrosis and Fibrotic Gene Expression in Pediatric and Adult Patients With Idiopathic Dilated Cardiomyopathy. J. Card. Fail. [Internet]. 2017 [cited 2023 Aug 31];23:314–324. Available from: https://pubmed.ncbi.nlm.nih.gov/27890770/

40. Miyamoto SD, Stauffer BL, Nakano S, Sobus R, Nunley K, Nelson P, Sucharov CC. Editor’s choice: Beta-adrenergic adaptation in paediatric idiopathic dilated cardiomyopathy. Eur. Heart J. [Internet]. 2014 [cited 2023 Aug 3];35:33. Available from: /pmc/articles/PMC3877432/

41. Patel MD, Mohan J, Schneider C, Bajpai G, Purevjav E, Canter CE, Towbin J, Bredemeyer A, Lavine KJ. Pediatric and adult dilated cardiomyopathy represent distinct pathological entities. JCI Insight [Internet]. 2017 [cited 2023 Aug 29];2. Available from: /pmc/articles/PMC5518561/

42. Kuszynski CA, Miller KA, Rizzino A. Influence of cell density and receptor number on the binding and distribution of cell surface epidermal growth factor receptors. In Vitro Cellular & Developmental Biology - Animal: Journal of the Society for In Vitro Biology [Internet]. 1993 [cited 2026 Mar 1];29:708–713. Available from: https://link.springer.com/article/10.1007/BF02631427

43. Seidel F, Scheibenbogen C, Heidecke H, Opgen-Rhein B, Pickardt T, Klingel K, Berger F, Messroghli D, Schubert S. Compensatory Upregulation of Anti-Beta-Adrenergic Receptor Antibody Levels Might Prevent Heart Failure Presentation in Pediatric Myocarditis. Front. Pediatr. [Internet]. 2022 [cited 2026 Mar 1];10. Available from: https://pubmed.ncbi.nlm.nih.gov/35573966/

44. Woo AYH, Song Y, Xiao RP, Zhu W. Biased β2-adrenoceptor signalling in heart failure: pathophysiology and drug discovery. Br. J. Pharmacol. [Internet]. 2014 [cited 2026 Mar 1];172:5444. Available from: https://pmc.ncbi.nlm.nih.gov/articles/PMC4667850/

45. Wadhawan R, Tseng YT, Stabila J, McGonnigal B, Sarkar S, Padbury JF. Regulation of cardiac beta 1-adrenergic receptor transcription during the developmental transition. Am. J. Physiol. Heart Circ. Physiol. [Internet]. 2003 [cited 2026 Mar 1];284. Available from: https://pubmed.ncbi.nlm.nih.gov/12742828/

46. Alabed S, Sabouni A, Al Dakhoul S, Bdaiwi Y. Beta-blockers for congestive heart failure in children. Cochrane Database Syst. Rev. [Internet]. 2020 [cited 2025 Sep 28];2020:CD007037. Available from: https://pmc.ncbi.nlm.nih.gov/articles/PMC7389334/

47. Patel MD, Mohan J, Schneider C, Bajpai G, Purevjav E, Canter CE, Towbin J, Bredemeyer A, Lavine KJ. Pediatric and adult dilated cardiomyopathy represent distinct pathological entities. JCI Insight [Internet]. 2017 [cited 2025 Sep 28];2:e94382. Available from: https://pmc.ncbi.nlm.nih.gov/articles/PMC5518561/

48. Rücker B, Almeida ME, Libermann TA, Zerbini LF, Wink MR, Sarkis JJF. E-NTPDases and ecto-5′-nucleotidase expression profile in rat heart left ventricle and the extracellular nucleotide hydrolysis by their nerve terminal endings. Life Sci. 2008;82:477–486.

49. Novitskaya T, Nishat S, Covarrubias R, Wheeler DG, Chepurko E, Bermeo-Blanco O, Xu Z, Baer B, He H, Moore SN, et al. Ectonucleoside triphosphate diphosphohydrolase-1 (CD39) impacts TGF-β1 responses: insights into cardiac fibrosis and function following myocardial infarction. Am. J. Physiol. Heart Circ. Physiol. [Internet]. 2022 [cited 2025 Oct 14];323:H1244. Available from: https://pmc.ncbi.nlm.nih.gov/articles/PMC9722260/

50. Zheng H, Yang Y, Han J, Jiang WH, Chen C, Wang MC, Gao R, Li S, Tian T, Wang J, et al. TMED3 promotes hepatocellular carcinoma progression via IL-11/STAT3 signaling. Sci. Rep. [Internet]. 2016 [cited 2025 Oct 14];6. Available from: https://pubmed.ncbi.nlm.nih.gov/27901021/

51. Cabral-Pacheco GA, Garza-Veloz I, Rosa CCD La, Ramirez-Acuña JM, Perez-Romero BA, Guerrero-Rodriguez JF, Martinez-Avila N, Martinez-Fierro ML. The roles of matrix metalloproteinases and their inhibitors in human diseases. Int. J. Mol. Sci. 2020;21:1–53.

52. Takawale A, Zhang P, Patel VB, Wang X, Oudit G, Kassiri Z. Tissue Inhibitor of Matrix Metalloproteinase-1 Promotes Myocardial Fibrosis by Mediating CD63-Integrin β1 Interaction. Hypertension [Internet]. 2017 [cited 2025 Oct 14];69:1092–1103. Available from: https://www.ahajournals.org/doi/10.1161/HYPERTENSIONAHA.117.09045

53. Kralisch S, Lossner U, Bluher M, Paschke R, Stumvoll M, Fasshauer M. Tissue inhibitor of metalloproteinase 1 expression and secretion are induced by beta-adrenergic stimulation in 3T3-L1 adipocytes. J. Endocrinol. [Internet]. 2006 [cited 2025 Oct 14];189:665–670. Available from: https://pubmed.ncbi.nlm.nih.gov/16731796/

54. Menon B, Singh M, Singh K. Matrix metalloproteinases mediate beta-adrenergic receptor-stimulated apoptosis in adult rat ventricular myocytes. Am. J. Physiol. Cell Physiol. [Internet]. 2005 [cited 2025 Oct 14];289. Available from: https://pubmed.ncbi.nlm.nih.gov/15728709/

55. Woulfe K, Lin Y, Li X, Mahaffey J, Sweet M, Taylor M, Mestroni L, Miyamoto S, Stauffer B, Sucharov C, et al. Age-specific changes in myofibril mechanics in pediatric dilated cardiomyopathy. J. Mol. Cell. Cardiol. [Internet]. 2018 [cited 2025 Dec 15];124:105–106. Available from: https://www.jmcc-online.com/action/showFullText?pii=S0022282818304681

